# A Novel Blended Hybrid Care Model for Maternal Mental Health: Cohort Study of Pregnant and Postpartum Patients

**DOI:** 10.64898/2026.03.07.26347860

**Authors:** Elombe Calvert, Kelly Chen, Khatiya Chelidze Moon, Margaret R. Emerson, Natalie Feldman, Catherine Lager, John Torous

**Author notes:** **Corresponding author, location of work & address for reprints:** John Torous, MD, MBI, Division of Digital Psychiatry, Beth Israel Deaconess Medical Center, 330 Brookline Avenue, Boston, MA 02115, USA.

## Abstract

**Background:** Perinatal mood and anxiety disorders are the most common complications of pregnancy. Given the limited mental health resources, there is a need for novel treatment approaches. Though smartphone applications can increase access to evidence-based care, recent research highlights notable limitations, including varying quality and unclear effectiveness. Blended hybrid care models, which integrate synchronous telehealth services with asynchronous modalities (such as mobile apps), have emerged as an alternative. This pilot study evaluates one such model, the Digital Clinic, to determine its potential to bridge this critical treatment gap and compare outcomes to that of non-peripartum patients in the clinic.

**Methods:** Pregnant and postpartum women referred for anxiety and depression received 8 weeks of synchronous, virtual, evidence-based CBT from a trained clinician. This treatment was complemented by the asynchronous use of the mindLAMP app, providing digital phenotyping, psychoeducation, and CBT skills, with the support of a Digital Navigator. The efficacy of the intervention was evaluated by comparing GAD-7 and PHQ-9 scores from intake to the end of treatment.

**Results:** This secondary analysis included 13 peripartum women from a larger sample of 224 clinic patients. At intake, they reported a mean PHQ-9 score of 9.4 (SD=3.9) and a mean GAD-7 score of 11.69 (SD=6.0). After 8 weeks, participants reported statistically significant decreases of 4.14 points on the GAD-7 (p<.01) and 3.92 points on the PHQ-9 (p<.01). Effect sizes for these reductions were 0.74 (95% CI: 0.20, 1.28) for GAD-7 and 1.10 (95% CI: 0.29, 1.90) for PHQ-9.

**Conclusion:** A novel blended hybrid care model, the Digital Clinic, was successful in reducing depression and anxiety among pregnant and postpartum women. This novel approach to maternal mental health shows promise for delivering accessible, effective, evidence-based care to peripartum patients in real-world settings. Future work should further validate its effectiveness with larger, more diverse patient populations with moderate to severe disease.

## Introduction

Perinatal mood and anxiety disorders (PMADs) are among the most common complications during pregnancy, significantly contributing to maternal morbidity and mortality [1]. The effects of maternal PMADs extend to infants, who may develop behavioral or learning difficulties [3]. Despite the considerable prevalence of perinatal depression (PD), access to quality, evidence-based mental health care remains limited. Recent data shows that only a fraction of patients with PD receive adequate treatment [4,5]. Consequently, there is a growing interest in effective, low-risk, and rapid-acting interventions to bridge the treatment gap and improve the quality of care for those affected.

While antidepressants are widely recognized as effective and supported by years of safety data for use during pregnancy and lactation, uncertainties remain regarding the risks associated with perinatal medication use. Due to ethical constraints on conducting research in pregnant and lactating populations, no double-blind, randomized controlled trials of antidepressants have been conducted in these groups. The United States Preventive Services Task Force (USPSTF) recommends psychotherapy as a first-line treatment for PD. While psychotherapy is an effective treatment for many, this treatment can take many weeks to show efficacy, and patient engagement in psychotherapy may be variable. There is a growing interest in novel approaches that augment PD care to address treatment gaps while minimizing potential risks.

Smartphone apps have great potential to improve the quality of care for perinatal patients. Apps geared towards improving the well-being of perinatal people are readily accessible. However, reviews of these apps suggest concerns about their data safety and variable efficacy [6,7]. Numerous studies have shown that apps are acceptable and of interest to both women with perinatal depression and their providers [8]. However, a 2022 review showed that despite the availability of apps for peripartum mental health, choosing the right one is an emerging challenge [9]. This challenge is compounded by recent reviews suggesting that the clinical quality of these apps is often low [10,11]. This is not to say that all apps focused on this population are unhelpful, as a 2024 meta-analysis of 176 randomized controlled trials of apps found an effect size of g=0.28 for depression [12], highlighting the potential of these apps to deliver effective care.

Given the limitations of standalone apps, there has been a move toward integrating them into hybrid models of care, which leverage a combination of in-person and app-based approaches to improve the scale and delivery of evidence-based treatments. Hybrid models of care combine self-guided app use with traditional clinical visits, offering a blend of in-person (or telehealth) care augmented by app use between sessions. Recent reviews suggest that hybrid models of care can be effective for depression even when offered over a shorter timeframe than traditional therapy. Thus, hybrid models of care may present a novel means to meet the USPSTF recommendations for offering evidence-based psychotherapy to prevent perinatal depression [13].

The hybrid care model is particularly well-suited for the peripartum population, whose demanding schedules necessitate flexible, remote options that reduce logistical barriers such as transportation and childcare required for traditional appointments. Perinatal individuals frequently experience distressing symptoms at various times throughout the day and night, underscoring the importance of interventions that are accessible at all hours. Such on-demand approaches provide support and accessibility that traditional care models, constrained to scheduled appointments during standard business hours, cannot offer. By addressing these unique temporal and logistical challenges, hybrid care models represent an innovative and practical solution tailored to the specific needs of the perinatal population, potentially enhancing engagement and improving clinical outcomes. Emerging research already suggests such hybrid care models are acceptable, but less is known about their clinical impact.

Our team has developed one such hybrid care model for depression and anxiety that has been well-studied across diverse populations. This model, called the Digital Clinic, offers 8 weeks of treatment with a blend of clinical sessions led by a therapist, self-guided app use with the mindLAMP smartphone app, self-tracking with the same mindLAMP smartphone app, and app engagement and troubleshooting support offered by a coach trained as a Digital Navigator. The model, run since 2018, has proven effective, and we have treated patients with PD in the clinic [11]. However, there has been no examination of the overall efficacy of this model of care for the peripartum population, nor have outcomes been compared with those of non-peripartum patients in the clinic. Thus, in this pilot analysis, we aim to evaluate the clinical effectiveness of our hybrid care model in addressing symptoms of anxiety and depression among peripartum women. Effectiveness will be measured by changes in depressive and anxiety symptoms from baseline to the completion of therapy. Additionally, as a secondary objective, we will compare the model’s effectiveness in peripartum women to its outcomes in non-peripartum patients.

## Methods

### Study Procedures

This secondary data analysis utilized peripartum patients, defined as pregnant or up to one year postpartum, who were referred to our hybrid care clinic model. Referred patients either had generalized anxiety disorder (GAD) or major depressive disorder (MDD) with peripartum onset. Our 8-week blended hybrid care model, based at the Beth Israel Deaconess Medical Center (BIDMC), is offered through clinician referral of patients from outpatient clinics across the Beth Israel Lahey Health (BILH) system. Our cohort of peripartum patients included those who participated in our hybrid care model between August 2022 and December 2024. Eligible participants were at least 18 years old, lived in Massachusetts, spoke English, and owned an Android or Apple smartphone. Once eligibility was confirmed, the participants met with a Digital Navigator who detailed the aspects of treatment and the role of mindLAMP in treatment and helped the patients install the app on their phone and log in and acquire informed consent. Once enrolled in the clinic, the Digital Navigator scheduled the patients’ first therapy appointment with a clinician and administered an electronic intake form designed to get a baseline assessment of clinical outcome measures. This form collected demographic data and responses to the Generalized Anxiety Disorder 7-item (GAD-7), Patient Health Questionnaire (PHQ-9), and the Sheehan Disability Scale. Enrolled patients received 8 weeks of evidence-based cognitive-behavioral therapy (CBT) from a trained clinician, following the Unified Protocol (UP) for Transdiagnostic Treatment of Emotional Disorders. This was complemented by using the mindLAMP mental health app, with support from a Digital Navigator who facilitated app use. Treatment was administered through weekly therapy sessions with the clinician, weekly check-ins with the Digital Navigator, and regular use of the mindLAMP app throughout the program. During check-ins, the Digital Navigator addressed any technical or motivational challenges related to the app and reviewed the patients’ active and passive digital data, providing insights and explanations. Notably, both therapy sessions and check-ins were conducted virtually via Zoom. A more comprehensive description of the hybrid care model’s methodology is available in our previous publication [14]. The app itself offers psychoeducation, self-assessments, CBT exercises, and real-time feedback, among other features. A series of screenshots of the app are shown below in Figure 1.

**Figure 1.**
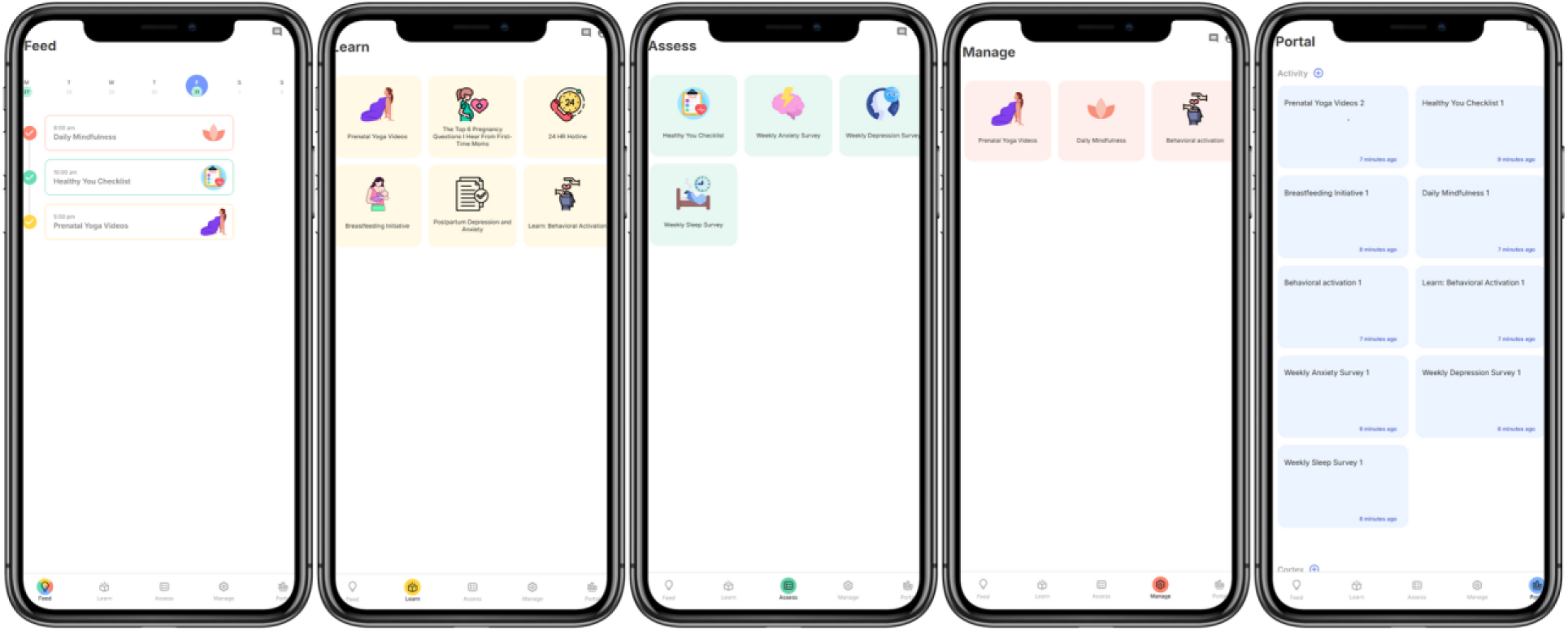
The mindLAMP App. This figure shows the different pages (Feed, Learn, Assess, Manage, Portal) of mindLAMP in addition to the resources, interventions, and data availability on the app accessible to users.

### Study Measures – Participant Features

#### Depressive symptoms

The 9-item Patient Health Questionnaire (PHQ-9) has been validated in prenatal depression, and [15] was used at baseline and each follow-up visit to assess the presence and severity of depressive symptoms over the preceding two weeks. Each of the nine items is scored on a scale from 0 (not at all) to 3 (nearly every day), yielding a total possible score of 0–27.

#### Anxiety symptoms

The 7-item Generalized Anxiety Disorder (GAD-7) has been validated in parental anxiety [16] was used at baseline and each follow-up visit to assess the presence and severity of anxiety symptoms over the preceding two weeks. Each of the seven items is scored on a scale from 0 (not at all) to 3 (nearly every day), yielding a total possible score of 0-21.

#### Self-efficacy at managing emotions

Self-efficacy in managing emotions was measured using the PROMIS Item Bank v1.0 – Self-Efficacy for Managing Emotions Short Form 8a [17]. Items are rated on a scale of 1 (I am not at all confident) to 5 (I am very confident), with a total possible score of 8-40. Higher scores indicated higher levels of self-efficacy for managing negative emotions.

#### Emotional self-awareness

Perceived emotional self-awareness was measured using a single question, “I feel as though I am aware of and in tune with emotions”, which was rated from 1 (I am not at all confident) to 5 (I am very confident). Higher scores indicated higher levels of self-awareness.

#### Perceived social support

The Multidimensional Scale of Perceived Social Support (MSPSS) [18] was used to measure levels of perceived social support. Items were rated on a scale of 1 (very strongly disagree) to 7 (very strongly agree), with a possible score range of 12-84. Higher scores indicated higher levels of perceived social support.

#### Functional Impairment

The Sheehan Disability Scale (SDS) [19], a 5-item assessment of impairment in 3 domains: work/school, social life, and family life. Three items assessing these three domains are rated from 0 (not at all) to 10 (extremely) and yield a total summed score of 0 (unimpaired) to 30 (highly impaired).

#### Self-Management Behaviors

The Partners in Health Scale (PIH) [20], a 12-item assessment with 3 subscales: knowledge, adherence and symptom management. Item responses were rated from 0 (Very little’, Never’, Not very well’) to 8 (A lot’, ‘Always’, ‘Very well’) and yield a total summed score 0 to 96; higher scores indicated more motivation.

#### Digital Literacy

The Digital Health Care Literacy Scale [21], a 3-item assessment, was used to measure digital health care literacy. The items are rated from 0 (strongly disagree) to 4 (strongly agree) and yield a total summed score 0 to 12. Scores that are closer to 12 indicate higher digital health literacy.

#### Demographic characteristics

Demographic data was collected at intake from patients and included age, race, and biological sex assigned at birth, educational attainment, and peripartum status while receiving care in the clinic.

#### mindLAMP engagement

Engagement with the mindLAMP app was evaluated through a set of usage metrics: the number of days daily surveys were completed (0–56), the number of weekly PHQ-9 and GAD-7 assessments completed (0–8), the number of weekly sleep surveys completed (0–8), the number of days active in the clinic (0–56) defined by the days on which patients engaged in any app-based activity during treatment and finally the total time spent using mindLAMP (with a minimum threshold of 10 hours).

### Statistical Analysis

Descriptive statistics, including means, standard deviations and frequencies, were used to summarize patient demographic characteristics, study measures at baseline and completion, and mindLAMP engagement metrics over the treatment period. To examine the potential efficacy of our hybrid model in decreasing symptoms of anxiety and depression in the peripartum cohort, we compared GAD-7 and PHQ-9 scores from baseline to the end of treatment by calculating the mean change for both measures. Independent t-tests were used to assess whether differences in scores between baseline and completion were statistically significant, using alpha levels of 0.05 or 0.01. Effect sizes (Cohen’s *d*) were computed to assess the magnitude of treatment impact. In addition, we evaluated the efficacy of our hybrid model in non-perinatal patients and compared their outcomes with those of the peripartum cohort.

## Results

### Baseline Participant Characteristics

Table 1 presents the demographic characteristics of the peripartum cohort. The mean participant age was 35.8 years (SD = 5.1), with 92% of the cohort being postpartum. Regarding racial composition, 69% identified as White, 15% as Black or African American, and 1% each as Asian and Middle Eastern/North African. Overall, the cohort was highly educated, with 54% holding a postgraduate degree, 31% having completed college, and 15% having pursued some college education.

**Table 1.** Baseline Characteristics of the Maternal Mental Health Cohort.

| Characteristic | Full Sample (N = 13) |  |
| --- | --- | --- |
|  | n /Mean<sup>1</sup> | % / SD<sup>2</sup> |
| Age | 35.8 | 5.1 |
| Sex |  |  |
| Female | 13 | 100 |
| Education |  |  |
| Some College | 2 | 15 |
| College Graduate | 4 | 31 |
| Postgraduate Degree | 7 | 54 |
| Race |  |  |
| Asian | 1 | 7.7 |
| Black or African American | 2 | 15 |
| Middle Eastern or North African | 1 | 7.7 |
| White | 9 | 69 |
| Maternal Status |  |  |
| Postpartum | 12 | 92 |
| Pregnant | 1 | 8.0 |

<sup>1</sup> Count (n); Mean<sup>2</sup> Percentage (%); SD

### Baseline and Completion Measures

Participants experienced a statistically significant reduction in both depressive (p<0.01) and anxiety (p<0.01) symptoms from baseline to completion (see Table 2). These decreases are illustrated in Figures 2 and 3, which show marked declines in symptom severity over the course of treatment. Significant improvements were also observed in self-efficacy for managing emotions (p<0.01) and self-management behaviors (p = 0.03). Notably, participants reported high levels of digital literacy at baseline (M = 11.8, SD = 0.6).

**Figure 2.**
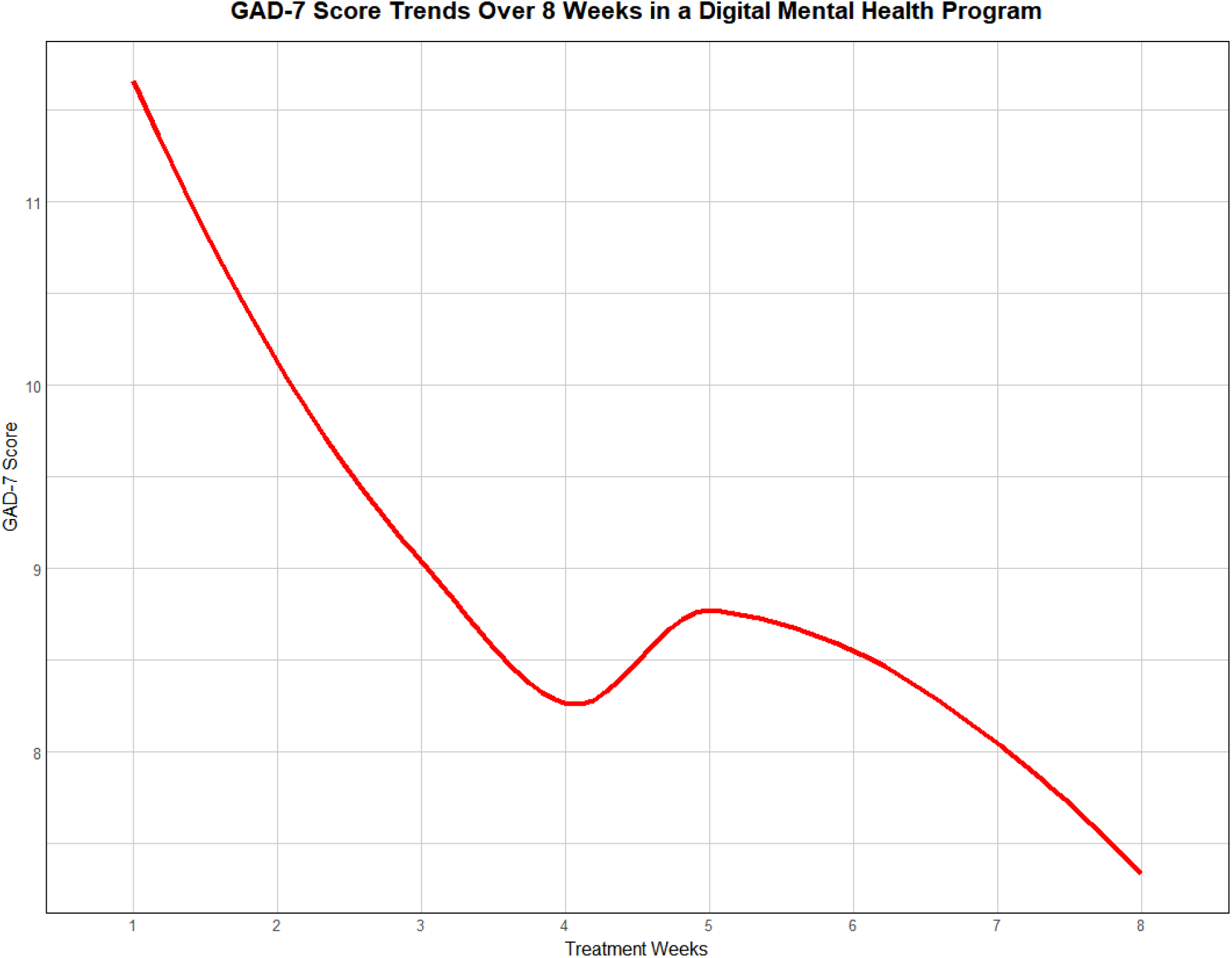
Trend in GAD-7 scores over an 8-week digital mental health program, illustrating a reduction in anxiety symptoms over time. Each point represents the average weekly score, highlighting fluctuations and overall improvement across the treatment duration.

**Figure 3.**
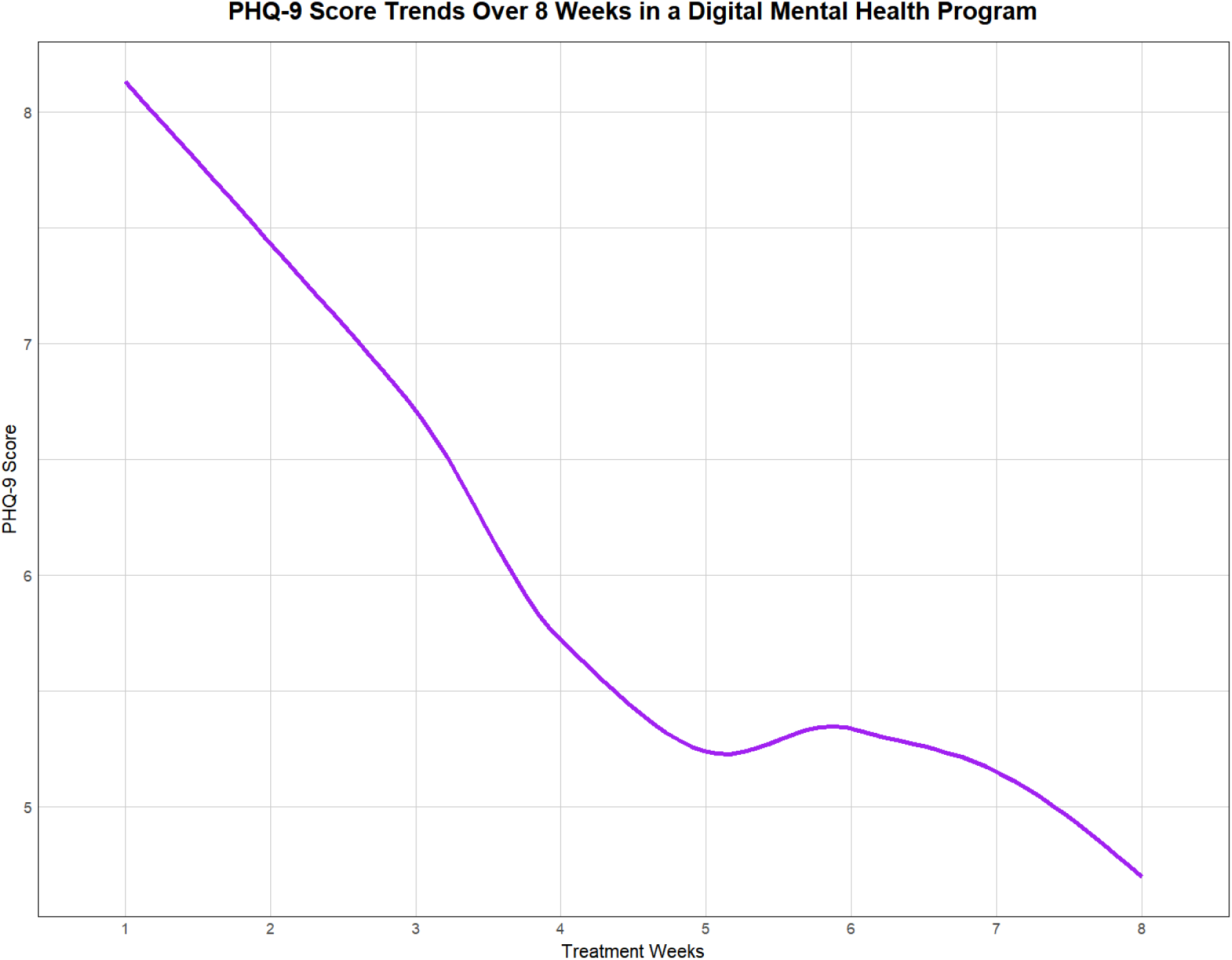
Trend in PHQ-9 scores over an 8-week digital mental health program, illustrating a reduction in anxiety symptoms over time. Each point represents the average weekly score, highlighting fluctuations and overall improvement across the treatment duration.

**Table 2.** Baseline and completion clinical scores for the maternal mental health cohort (n=13)

| Characteristic | Baseline Scores |  | Completion Scores |  | p-value |
| --- | --- | --- | --- | --- | --- |
|  | *M*<sup>1</sup> | *SD*<sup>2</sup> | *M*<sup>1</sup> | *SD*<sup>2</sup> |  |
| Anxiety Symptoms (GAD-7) | 11.7 | 6.0 | 7.5 | 2.8 | <0.01 |
| Depressive Symptoms (PHQ-9) | 9.38 | 3.9 | 5.5 | 3.1 | <0.01 |
| Functional Impairment | 18.0 | 9.0 | 12.0 | 10.0 | 0.12 |
| Perceived Social Support | 71.0 | 12.0 | 70.0 | 16.0 | 0.80 |
| Self-Efficacy at Managing Emotions | 19.3 | 3.5 | 27.0 | 8.0 | <0.01 |
| Self-Management Behaviors | 84.0 | 16.0 | 94.0 | 14.0 | 0.03 |
| Emotional Self-Awareness | 3.8 | 0.8 | 3.7 | 0.9 | >0.9 |
| Digital Health Literacy Skills | 11.8 | 0.6 | 11.7 | 0.9 | 0.56 |

<sup>1</sup> Mean (*M*)<sup>2</sup> Standard deviation (*SD*)
*Note.* **Depressive symptoms**, Patient Health Questionnaire-9 [score range: 0–27]. **Anxiety symptoms**, Generalized Anxiety Disorder-7 [score range: 0–21]. **Functional Impairment**, Sheehan Disability Scale [score range: 0–30]. **Perceived Social Support**, Multidimensional Scale of Perceived Social Support [score range: 12–84]. **Self-Efficacy at Managing Emotions**, PROMIS Item Bank v1.0 - Self-Efficacy for Chronic Conditions – Managing Emotions – Short Form 8a [score range: 8–40]. **Emotional Self-Awareness**, Emotional Self-Awareness Assessment [score range: 1–5]. **Digital Literacy**, Digital Health Care Literacy Scale [score range: 0–12]. **Self-Management Behaviors**, Partners in Health Scale [score range: 0–96].

### Cohort Mean Change and Effect Sizes

Tables 3 and 4 summarize changes in GAD-7 and PHQ-9 scores from baseline to the end of treatment, as well as the effect sizes of the hybrid clinic model for reducing depressive and anxiety symptoms among peripartum and non-peripartum patients. For depression symptoms in peripartum women, the hybrid model achieved a mean PHQ-9 reduction of –3.92 (SD = 5.06) with a large effect size of 1.10 (95% CI: 0.29, 1.28), compared to a mean PHQ-9 reduction of –3.63 (SD = 5.20) and a medium effect size of 0.63 (95% CI: 0.48, 0.77) among non-peripartum patients. Regarding anxiety symptoms, peripartum patients experienced a mean GAD-7 reduction of –4.15 (SD = 4.70) with a medium effect size of 0.74 (95% CI: 0.20, 1.28), whereas non-peripartum patients showed a mean GAD-7 reduction of –3.12 (SD = 5.00) with a medium effect size of 0.63 (95% CI: 0.48, 0.77).

**Table 3.** Effect Size of the Digital Clinic for Reducing Symptoms of Anxiety and Depression in Pregnant and Postpartum Patients (n=13).

| Score | Baseline,<br>mean (SD) | Follow-up,<br>mean (SD) | Change,<br>mean (SD) | P value<sup>1</sup> | Effect size<sup>2</sup> (95% CI) |
| --- | --- | --- | --- | --- | --- |
| **PHQ-9<sup>a</sup>** | 9.38 (3.9) | 5.46 (3.1) | -3.92 (4.0) | <0.01 | 1.10 (0.29, 1.90) |
| **GAD-7<sup>b</sup>** | 11.69 (6.0) | 7.53 (2.8) | -4.15 (4.7) | <0.01 | 0.74 (0.20, 1.28) |

<sup>a</sup>PHQ-9: 9-item Patient Health Questionnaire
<sup>b</sup>GAD-7: Generalized Anxiety Disorder-7
<sup>1</sup>T-test
<sup>2</sup>Cohen's d

**Table 4.**
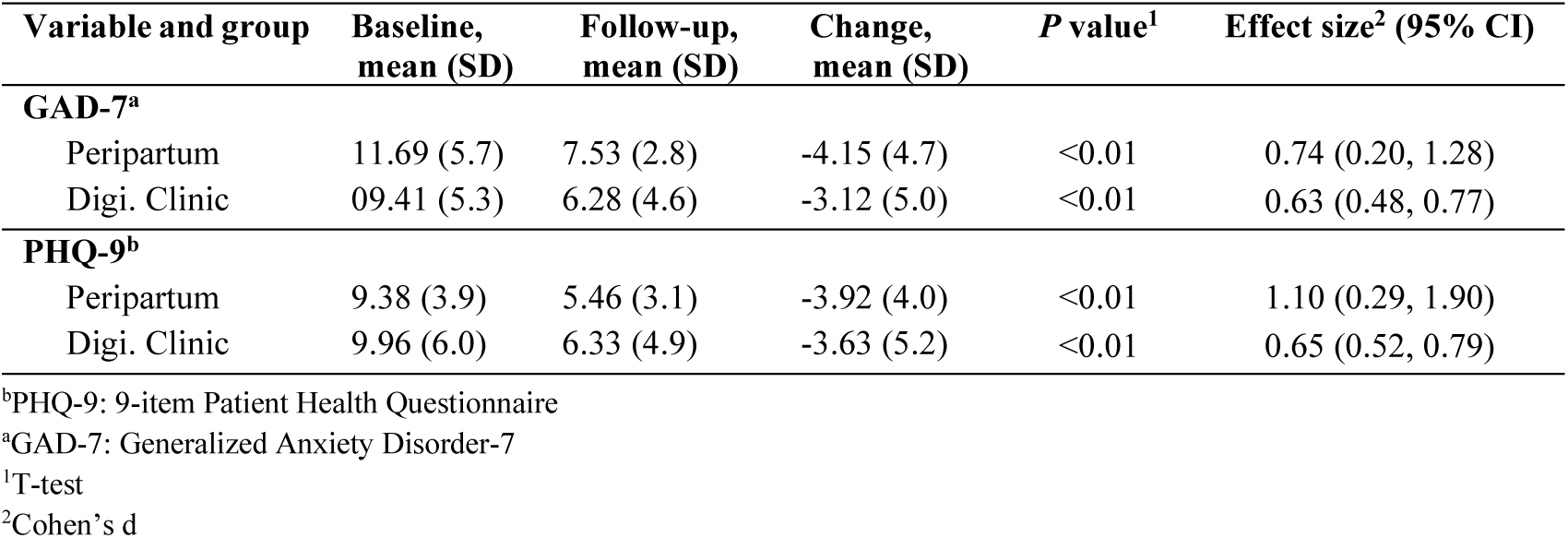
Differences and changes in GAD-7 and PHQ-9 scores at baseline and different follow-up intervals for peripartum patients (n=13) compared to non-partum patients (n=224) in our hybrid model.

## Discussion

Our preliminary results show that the hybrid care model demonstrated significant effectiveness in reducing symptoms of depression and anxiety among peripartum women. Additionally, the model proved to be more effective in the peripartum population compared to the general clinic population, achieving larger reductions in symptoms. These pilot results demonstrate that a blended, hybrid clinic model employing an open-source smartphone app has potential to improve care for PMADs. While there has been related past research, the proprietary nature of those efforts [22] limits scalable science, reproducible research, and follow-up implementation efforts. Our approach and results differ as they offer a foundation for broad, collaborative, and scalable efforts ready for implementation. By combining aspects of smartphone measurements, this model can aid in screening [13] and informed data use by directly connecting women needing care [23]. The hybrid nature of this approach addresses challenges posed by inadequate access to quality, evidence-based care for peripartum women [24] and highlights how digital mental health interventions are well suited to address this gap [25].

While hybrid care models can vary in the length of treatment offered, the shorter eight-week duration of this treatment has implications for treating PMAD. Clear evidence has shown that treating PMAD, in particular, has an immediate positive impact on infants, and delay can have implications for bonding [26]. During pregnancy, minimizing the effects of both depression and medication exposure on the woman and developing fetus is urgent, creating a need for fast-acting treatments with limited risks. The eight-week time frame of this treatment is also in line with the duration that most antidepressants may take to become effective, offering a viable alternative to antidepressant treatment as well as traditional psychotherapy care, which may take longer than eight weeks to be effective. The virtual nature of this treatment can also help provide more flexibility to patients with PMADs.

Our results also suggest one possible mechanism of action. As shown in Table 2, self-efficacy at managing emotions scores improved across the treatment. Given this treatment’s focus on using the app to practice skills related to emotional regulation and tracking emotions daily, the asynchronous aspects of this approach may impart these additional benefits beyond the clinical benefit of weekly virtual sessions with a therapist. It may also reflect the support of the Digital Navigator to help drive app use and the clinical intervention of the app into care during the weekly therapy sessions [27].

Compared to our clinic’s outcomes in patients referred broadly from primary care, outcomes for perinatal patients were superior for both anxiety and depression. While our study does not allow us to assess why, potential mechanisms could include perinatal patients’ being more engaged with the asynchronous aspects of this treatment (i.e., the app) or experiencing more behavioral symptoms (e.g. sleep disturbances) that this treatment approach can quantify, increasing potential for self-help behaviors as well as the engagement of the treatment team. For example, the digital phenotyping aspects of the app can help capture the sleep patterns of new mothers and ensure that data is routed to the clinical team for consideration in treatment plans.

This pilot study has several limitations. The sample size is small, highly digitally literate, predominantly college-educated, and Caucasian, which may limit the generalizability of the results. While these preliminary results are promising, future studies could examine the effects of the model on patients with moderate-to-severe disease, as our participants had low PHQ-9 and GAD-7 scores at baseline. However, these limitations can be addressed in a larger study that can build off the feasibility and promising pilot results while also adapting aspects of the treatment for underserved communities [27, 28] and offering digital literacy support to those who need it to partake [29, 30].

## Conclusion

The Digital Clinic, a novel blended hybrid care model, has demonstrated success in reducing depression and anxiety among pregnant and postpartum women. This approach to maternal mental health shows promise for delivering accessible, effective, and evidence-based care to peripartum patients in real-world settings. Future work should further validate its effectiveness with larger, more diverse patient populations with moderate to severe disease.

## Data Availability

All data produced in the present study are available upon reasonable request to the authors.

## List of Abbreviations

CBT: Cognitive-Behavioral Therapy
GAD-7: Generalized Anxiety Disorder 7-item
PHQ-9: 9-item Patient Health Questionnaire
PMADs: Perinatal mood and anxiety disorders
PD: perinatal depression
USPSTF: United States Preventive Services Task Force
GAD: generalized anxiety disorder
MDD: major depressive disorder
BIDMC: Beth Israel Deaconess Medical Center
BILH: Beth Israel Lahey Health
UP: Unified Protocol
MSPSS: Multidimensional Scale of Perceived Social Support
SDS: Sheehan Disability Scale
PIH: Partners in Health Scale
SD: Standard Deviation
CI: Confidence Interval

## Declarations

### Ethics approval and consent to participate

The operations of our hybrid model and the use of program data for research were reviewed and approved by the Beth Israel Deaconess Medical Center (BIDMC) Institutional Review Board (IRB) as part of a quality improvement project. As part of the enrollment process, patients provided informed consent during intake. During this process, they met with a Digital Navigator who explained the treatment, the structure of the model, and the collection of digital phenotyping data through mindLAMP, as well as how this data would be used both during and after treatment. Patients then signed an Informed Consent & Acknowledgement of Services form, allowing their data to be de-identified and used for research purposes.

### Consent for publication

Not Applicable

### Availability of data and materials

The datasets generated and/or analyzed during the current study are not publicly available as they contain identifiable patient information, however, an anonymized version can be made available upon request.

### Competing interests

JT is an advisor to Boehringer Ingelheim outside of the submitted work.

### Funding

No funding was received for this study

### Authors’ contributions

All authors contributed equally, helped write, reviewed, and approved the final manuscript.

## Acknowledgments

Not applicable

## Notes

### Funding Statement

This study did not receive any funding.

### Author Declarations

Ethics committee/IRB of Beth Israel Medical Deaconess Center gave ethical approval for this work.

